# Characterization of the redox status of cancer patients through the d-ROMs and BAP test and correlation of these parameters with blood variables

**DOI:** 10.1101/2023.01.26.23285045

**Authors:** Clarissa Aires de Oliveira, Eugenio Luigi Iorio, Lara Ferreira Paraiso, Foued Salmen Espindola

**Affiliations:** Institute of Biotechnology, Federal University of Uberlandia, Uberlandia, MG, Brazil; International Observatory of Oxidative Stress. 84127, Salerno. Italy

**Keywords:** oxidative stress, biological antioxidant potential, reactive oxygen metabolites, cancer

## Abstract

Oxidative distress and inflammation are common biochemical disorders in individuals with cancer. The measurement of oxidative stress in oncology can be useful in clinical practice to monitor the effectiveness of therapy and unwanted effects of the treatment. Thus, the aim of the present study was to evaluate the redox status through the reactive oxygen metabolites (d-ROMs) and biological antioxidant potential (BAP) tests and investigate the correlations of these parameters with blood variables in cancer patients. This is an observational, retrospective study of analysis of medical records of patients evaluated the period from 2018 to 2020 in an integrative medicine center. The inclusion criteria were individuals of both sexes, over 18 years of age, diagnosed with cancer who performed the d-ROMs and BAP test in the same period of blood analysis. Following the inclusion criteria, the final sample of the study were 57 individuals, 60% were woman and 40% were men. The evaluation of redox state showed that the d-ROMs were high (420.2 ± 112.1 U CARR) in total sample and higher in women compared to male (p < 0.01) and BAP tests were normal (2332 ± 812 μmol/l). The oxidative parameters, d-ROMs and OSI, was correlated positively with BAP, red cell distribution width (only d-ROMs), platelets (Plt), C-reactive protein (CRP), erythrocyte sedimentation (ESR) and negatively with hemoglobin (Hb) and mean corpuscular hemoglobin (MCH). Regarding the antioxidant potential index, BAP/dROMs, were correlated positively with Hb and serum albumin (HAS) and negatively correlated with Plt, CRP and ESR. The study shows that redox status of an individual with cancer is altered, and it is possible to monitor this system in clinical practice through d-ROMs and BAP test. These parameters, in addition to being suitable for assessing oxidative stress, were correlated with parameters predictors of inflammation.

## INTRODUCTION

Oxidative stress is a variant of the biological phenomenon called “stress” and is the adaptative response enabled by any living organism to face a series of physical, chemical, and biological stressors and, therefore, to survive (1). In the last years, new terms were introduced to classify subforms of oxidative stress and to conceptually introduce intensity scales ranging from physiological oxidative stress (eustress) to excessive and toxic oxidative burden (distress) (2–5).

The distress is a condition in which the redox system is unable to manage the exchange of reducing equivalents and the consequences are deleterious and can be of functional nature (e.g., a signaling or defense mechanisms impairment) and/or structural nature (e.g., a damage to proteins, lipids, nucleic acids, etc.) (2,3,6–9). On this basis, oxidative distress is involved in the pathogenesis and progression of several diseases including cancer (10–14).

Oxidative distress is not a disease, in the classical sense of the term, but rather an emerging health risk factor which, unfortunately, does not give rise to any specific clinical picture; it’s symptoms or signs are in fact obscured by the underlying diseases with which it is associated (7,15–18) and can only be diagnosed through specific tests.

Several approaches are currently available to assess the redox function or state of an individual, ranging from *ex vivo* free radical determination through electron spin magnetic resonance to *in vivo* imaging methods. Unfortunately, most of these approaches are not yet applicable for clinical examination because of the instability of many reactive species and the need for expensive equipment (19). This made it difficult for many years to translate the enormous achievements of science in the field of redox phenomena into clinical practice.

However, over the last three decades different approaches have become available (20–23) with the aim to measure oxidative stress in the clinical practice. In this sense, there are some tests, based on the principle of photometry, that show great cost/benefit, highly sensitivity and specificity (24,25). The reactive oxygen metabolites (d-ROMs) and biological antioxidant potential (BAP) test are based on this principle and are validated methods for measuring the pro-oxidant component and antioxidant capacity in plasma respectively (26–29).

By combining the results of d-ROMs and BAP test (alone or as oxidative stress indexes, i.e., OSI or BAP/d-ROMs ratios) it is possible to have suitable information about the redox status of the whole organism (26,28,30). In fact, both the d-ROMs and BAP tests have been shown to meet almost all the criteria of an ideal biomarker of oxidative stress for clinical practice (31–36) and have also proven useful in the evaluation of redox status in patients at risk for cancer or suffering from cancer, before and after antineoplastic treatment (37–41).

The possibility of globally measuring the redox function, through a careful evaluation of both the oxidant state and the reducing / antioxidant state, opens promising scenarios in the oncology field. In fact, the discovery of alteration oxidative stress biomarkers during a periodic health checkup, can be a valid clue to start more specific investigations aimed at highlighting neoplastic processes hitherto silent before they emerge clinically. For example, several epidemiological studies of oxidative stress markers in cancer, proved that high d-ROMs values are associated with prognosis of lung, colorectal, follicular lymphoma, and breast cancer (37,41–45).

Despite being reliable and widely used tests in several countries (24,36,46–49), d-ROMs and BAP tests have not yet been used in the Brazilian population. In addition, there is little evidence of the correlation of these parameters with blood variables that predict inflammation. Thus, the aim of the present study was to evaluate the redox status through the d-ROMs and the BAP test of Brazilian individuals with cancer and investigate the correlations of these parameters with hematological and biochemical variables.

## MATERIALS AND METHODS

### Population

The study was approved by the Human Research Ethics Committee (nº 5.671.038/2022) of the UNA University Center. This is an observational, retrospective study, analyzing the medical records of patients evaluated in the period 2018 to 2020 at the Conceito Saúde Clinic (CSC) located in Uberlandia - Minas Gerais, Brazil. The CSC is a center which specializes in integrative medicine where the patient’s diagnostic and the therapeutic follow-up includes a detailed investigation of the individual’s biochemical, cellular and physiological processes, which includes the assessment of the redox status.

Thus, in the established period, more than 600 d-ROMs and BAP tests were performed at the center, and the inclusion criteria for selecting sample in the present study were: 1) individuals of both sexes, over 18 years of age and diagnosed with cancer (through laboratory diagnostic methods); 2) patients who performed hematological and biochemical tests in the same period as the d-ROMs and BAP test. Thus, following the inclusion criteria established the final sample of the study consisted of 57 individuals (Fig 1).

**Fig 1.**
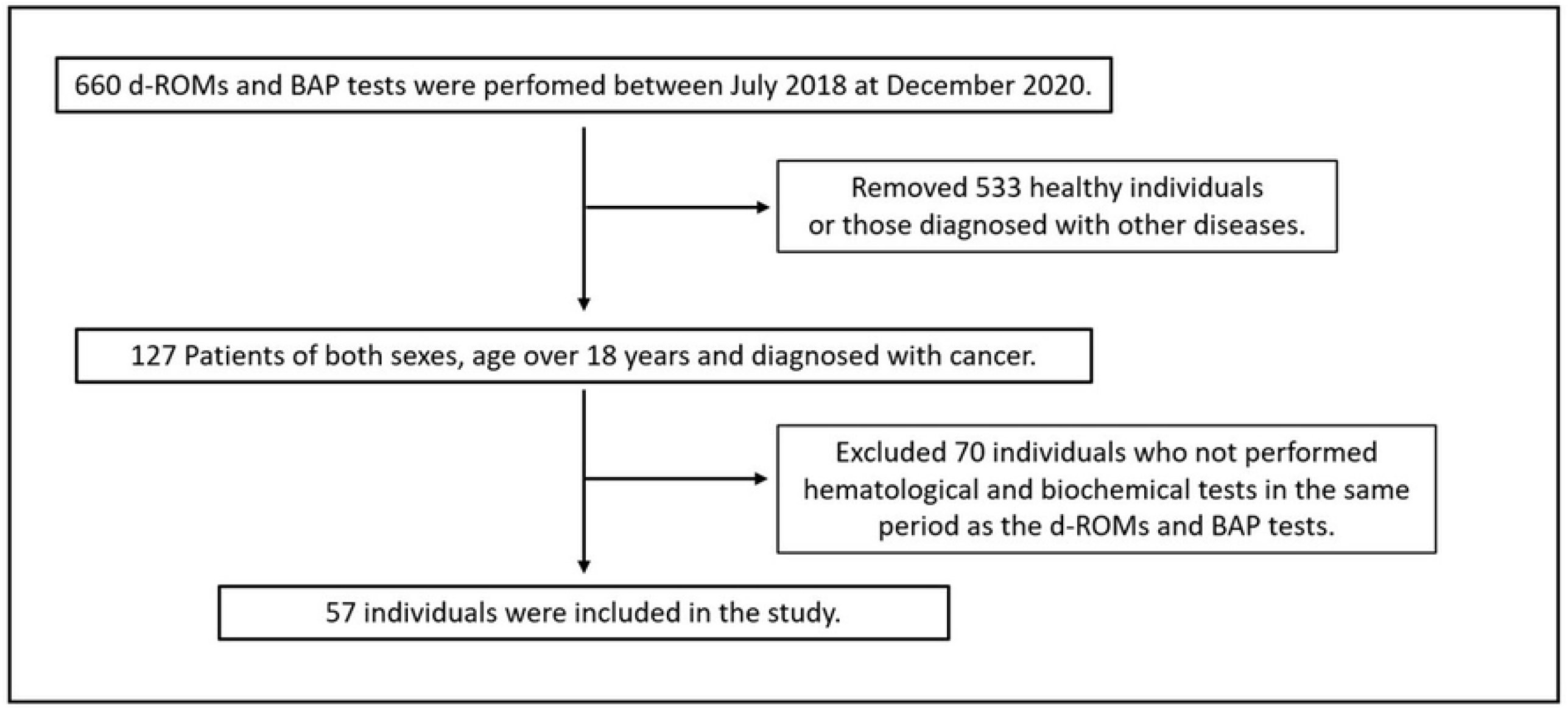
Study sample selection criteria. Abbreviations: Reactive oxygen metabolites (d-ROMs), biological antioxidant potential (BAP)

### Blood collections and determination of hematological and biochemical variables

After fasting for 8–12 h blood samples were collected in evacuated tubes (Vacutainer, Becton Dickinson, Juiz de Fora, Brazil) containing EDTA for hematological evaluation and without anticoagulant for biochemical determinations. The collection was carried out in the CSC and transported properly refrigerated (temperatures between 0 and 4°C) to the laboratory LABORMED, in Uberlandia, MG, Brazil where the analysis was performed.

Routine hematological variables, including erythrocytes (RBC), total leukocytes (Leu) and platelets (Plt) counts, hemoglobin (Hb), hematocrit (Ht), mean corpuscular volume (MCV), mean corpuscular hemoglobin (MCH), mean corpuscular hemoglobin concentration (MCHC), red cell distribution width (RDW) and erythrocyte sedimentation rate (ESR) were obtained in an automated hematology analyzer Sysmex XE 2100 (Sysmex Corporation, Kobe, Japan). The serum sample was used for the determination of biochemical parameters. Using the calorimetric method, the variable human serum albumin (HSA) was obtained. C-reactive protein (CRP) was obtained by turbidimetry method and direct bilirubin (DB) obtained by the colorimetric diazo method. All biochemical variables were determined with an automated biochemical analyzer Cobas 6000 (Roche, Germany).

### Measurements of d-ROMs and BAP test

The reactive oxygen metabolites (d-ROMs) test is based on this principle which exploits the chromogenic properties of an aromatic amine, i.e., N, N-diethylparaphenylenediamine. This compound, in its “basal” state, dissolved in water, gives rise to a colorless, transparent solution; however, when oxidizing species are added, it easily releases its electrons, causing the color of the solution to turn pink-red. Setting the wavelength of a photometer at 505 nm, it is possible to convert this oxidizing “power” into a number, through which the result of the d-ROMs test can be expressed (29,50).

For the measurement of d-ROMs, 10 μl of fresh plasma and 20 μl of chromogenic substrate (reagent R1) were mixed with 1 ml of buffer pH 4.8 (reagent R2). All the solutions were delicately mixed for 10 seconds and were read in spectrophotometer, by measuring their absorbance at 505 nm. Results were expressed in arbitrary units with normal values ranging from 250-300 U CARR, values above 300 indicate oxidative stress. The BAP test exploits the chromogenic properties of the iron complex of isocyanate derivative. A water solution of such ferric ions (Fe^3+^) complex is red; however, when antioxidant species are added, the Fe^3+^ions capture the electrons they donate and, transforming into ferrous ions (Fe^2+^), cause the breakdown of the colored complex, so that the solution becomes less red, tending to transparent. The more intense the observed color variation (from red-pink to colorless), the greater the “anti-oxidant power” of the biological sample examined and, consequently, the greater the capacity of the redox system to make available antioxidant species. A photometer will convert this antioxidant “power” into a number, through which the result of the BAP test will be expressed (26).

For the measurement of BAP, 10 μl of plasma were dissolved in 1 ml of a solution previously prepared by dissolving 50 μl of a thiocyanate derivative (R1 reagent) in 950 μl of a solution of ferric chloride (reagent R2). The solution was mixed by inversion for 10 seconds and the reduction of ferric ion was quantitated by measuring the change in absorbance at 505 nm, indicating the antioxidant activity of the serum sample. Values ≥2200 μmol/l were considered optimal; values below indicate oxidative stress due to low level of antioxidant activity/level (26).

The d-ROMs and BAP tests were performed using the FRAS 4 analyzer (Health & Diagnostics Limited Co., Parma, Italy) and the reagents for both tests were purchased from Wismerll Co., Ltd (Tokyo, Japan).

### Calculation of the Oxidative stress index (OSI) and BAP/dROMs

The ratio of d-ROMs to BAP gives the OSI (51), this ratio has been proven to be clinically relevant for evaluating the severity of certain diseases and since the shift of oxidative/antioxidative balance toward the oxidative side, is considered to represent oxidative stress (52,53).

In order to highlight the differences between antioxidant and pro-oxidant potential the ratio between BAP and d-ROMs was evaluated (BAP/dROMs) (54–56).

### Statistical analysis

Data analysis was performed by using IBM SPSS statistics version 21.0 (SPSS, Chicago, IL). Data distribution was performed using the Shapiro Wilk test. Differences between groups were calculated by unparied t test (for normality variables) or Mann Whitney test (for nonparametric variables). Correlations between variables tested were assessed by Spearman’s correlation coefficient. A value of p ≤ 0.05 was taken to be statistically significant.

## Results

The characterization of the sample is presented in table 1, with 34 (60%) were female and 23 (40%) male patients. Breast cancers were most frequent, followed by prostate, lung, bowel and melanoma of skin.

**Table 1.**
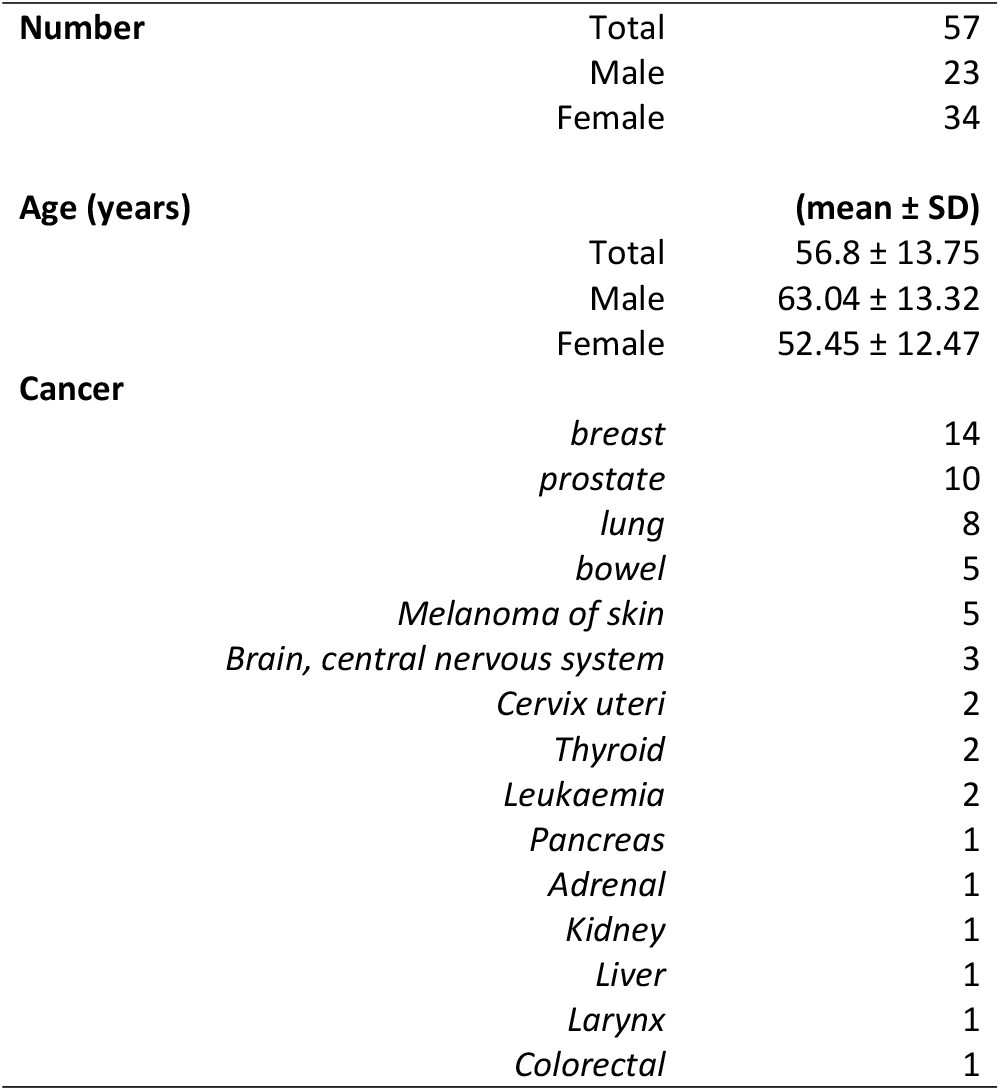
Baseline characteristics of the study population.

Fig 2 presents the results of the assessment of the redox status of patients. In total sample the mean of the d-ROMs were 420.2 ± 112.1 (U CARR), BAP test 2332 ± 812 (μmol/l), OSI ratio 0.16 ± 0.05 and BAP/dROMs ratio 6.02 ± 2.88. Comparison of redox tests between the sexes revealed that d-ROMs values were higher in women (p < 0.01).

**Fig. 2.**
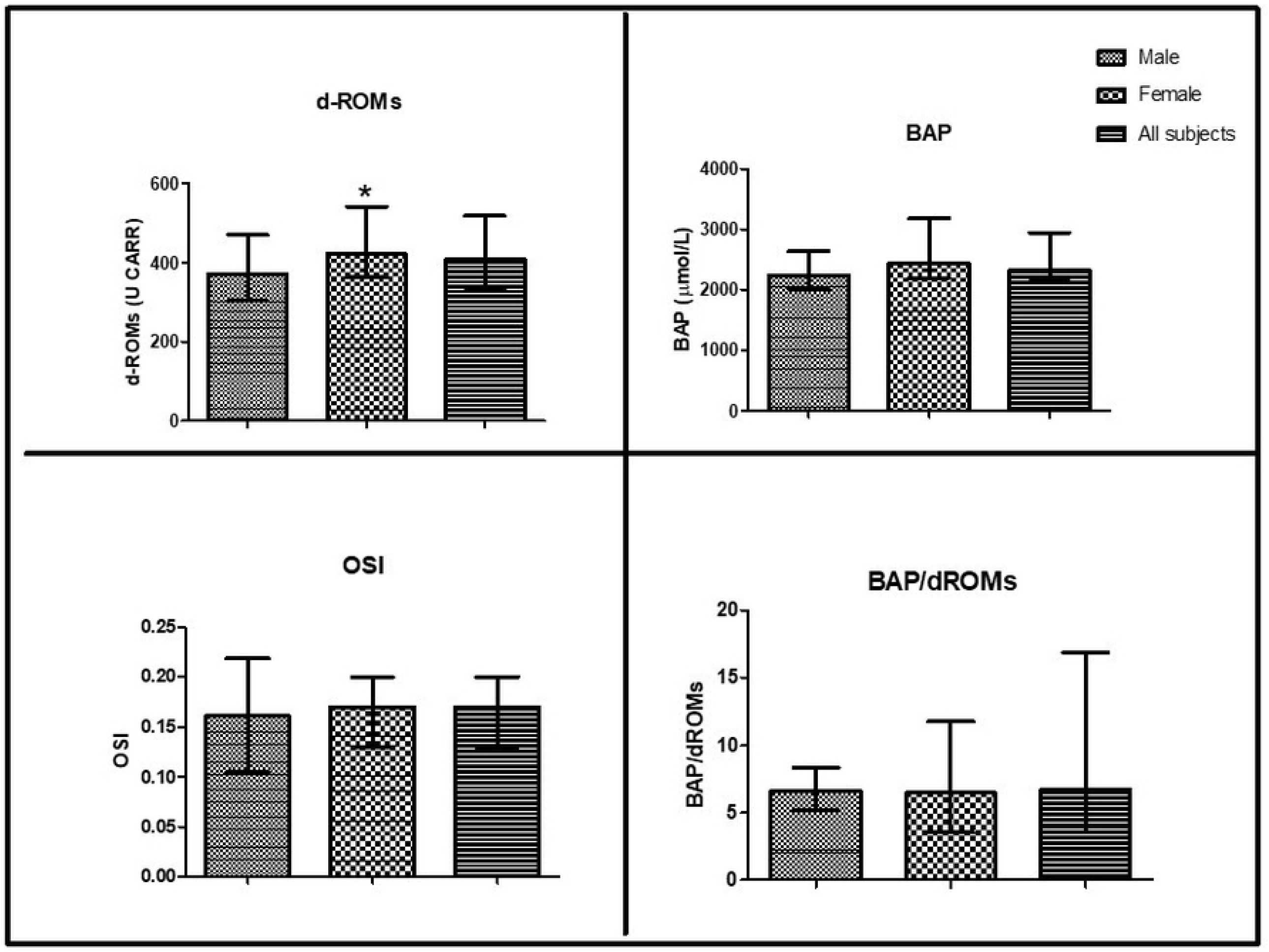
Characterization of the subjects’ redox system using the d-ROMs and BAP test. * Statistically significant difference (p< 0.01) when compared with same variable male.

Table 2 presents the characterization of the evaluation variables of the hematological and biochemical profile that were used to test correlations with the redox state parameters showed in table 3.

**Table 2.** Characterization of the hematological and biochemical profile of study subjects (N=57).

|  | Male | Female | All subjects | RV |
| --- | --- | --- | --- | --- |
| **Rbc ($10^6/\text{mm}^3$)** | 4.26 ± 0.92 | 4.2 ± 0.51 | (4.31 ± 0.75) | 4.3 – 5.7 |
| **Hb (g/dL)** | 12.8 ± 2.84 | (12.8 ± 1.4) | (12.9 ± 2.10) | 13.5 – 17.5 |
| **Ht (%)** | 37.99 ± 7.9 | 37.61 ± 3.91 | (38.6 ± 6.75) | 39.0 – 50.0 |
| **MCV (fL)** | 89.51 ± 5.53 | 89.65 ± 5.86 | 89.59 ± 5.67 | 81.0 – 95.0 |
| **MCH (pg)** | 30.05 ± 2.17 | 30.02 ± 2.46 | 30.03 ± 2.33 | 26.0 – 34.0 |
| **MCHC (g/dL)** | 33.41 ± 1.45 | 33.49 ± 1.22 | 33.46 ± 1.31 | 29.0 – 37.0 |
| **RDW (%)** | (13.15 ± 2.85) | 14.06 ± 1.69 | (13.4 ± 2.4) | 12.0 – 15.0 |
| **Leu ($\text{mm}^3$)** | 6.420 ± 2.613 | (5.000 ± 2.440) | (5.23 ± 2.76) | 3.500 – 10.500 |
| **Plt ($\text{mm}^3$)** | 218.8 ± 77.49 | 232.5 ± 91.90 | (211.0 ± 92.8) | 135.000 - 450.000 |
| **CRP (mg/L)** | (4.44 ± 19.06) | (1.47 ± 5.9) | (1.69 ± 9.84) | less than 1.0 |
| **HAS (g/dL)** | 4.38 ± 0.56 | 4.28 ± 0.53 | 4.32 ± 0.54 | 3.5 a 5.2 |
| **DB (mg/dL)** | 0.53 ± 0.25 | (0.43 ± 0.19) | (0.44 ± 0.33) | less than equal to 0.30 |
| **ESR (mm)** | (18.00 ± 43.00) | 32.9 ± 30.07 | (23.5 ± 41.75) | Male: 0 – 15<br>Female: 0 – 20 |

Data were expressed as mean ± SD or median and interquartile range ( ). \* Statistically significant difference ( $p < 0.05$ ) when compared with same variable male.
Abbreviations: Reference value (RV), erythrocytes (RBC), hemoglobin (Hb), hematocrit (Ht), mean corpuscular volume (MCV), mean corpuscular hemoglobin (MCH), mean corpuscular hemoglobin concentration (MCHC), red cell distribution width (RDW), total leukocytes (Leu), platelets (Plt), C-reactive protein (CRP), human serum albumin (HSA), direct bilirubin (DB) and erythrocyte sedimentation rate (ESR).

**Table 3.**
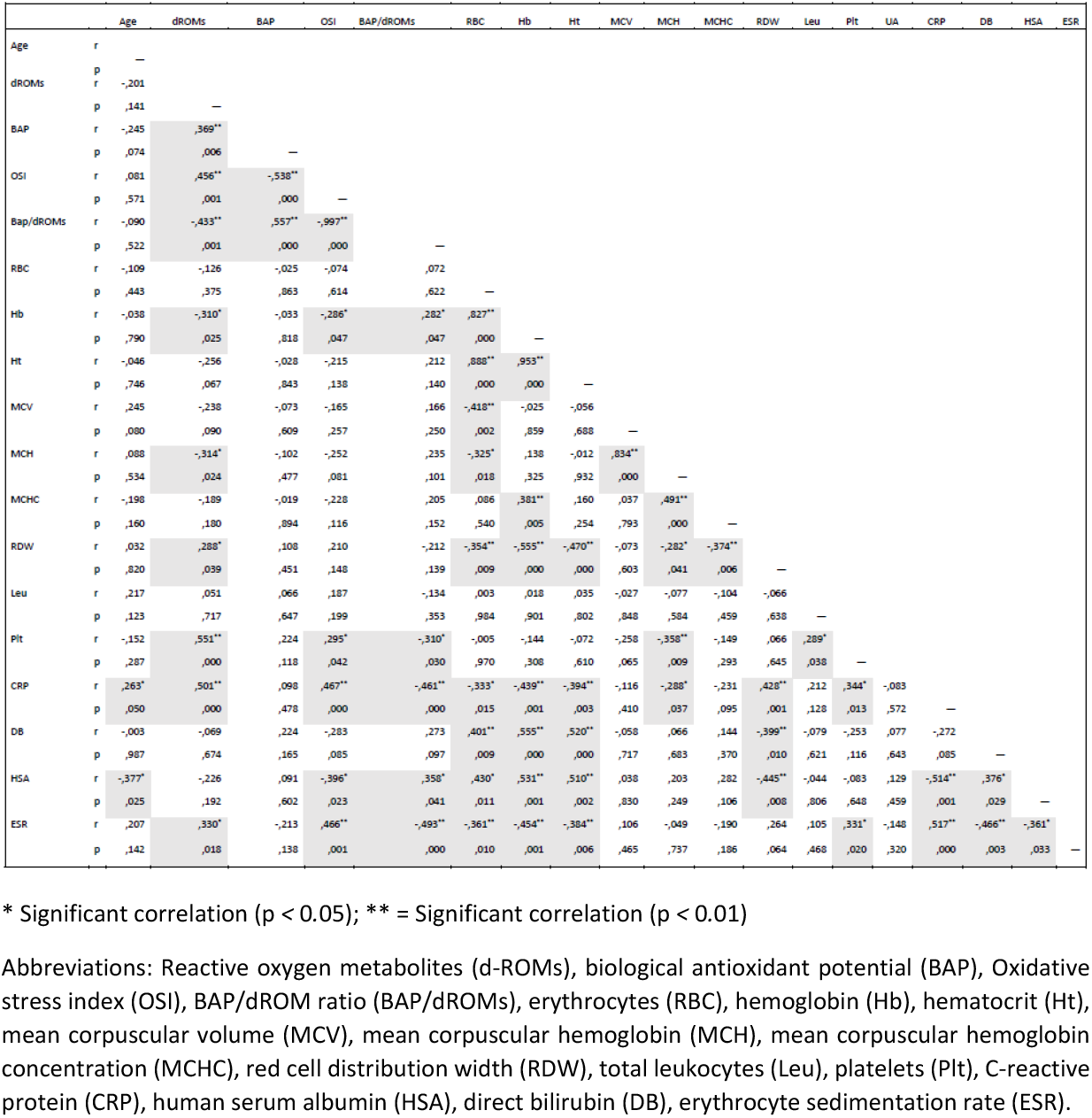
Spearman Correlation between pairs of all subjects’ variables (N=57).

The Spearman correlation of all blood variables with the parameters of evaluation of the redox status was performed. It was found that d-ROMs was correlated positively with BAP, RDW, Plt, CRP, ESR and negatively with Hb and MCH. Regarding the OSI was positively correlated with Plt, CRP and ESR; negatively correlated with Hb and HAS. Ultimately BAP/dROMs correlated positively with Hb and HAS and negatively correlated with Plt, CRP and ESR (Table 3).

Regarding blood parameters, RDW, Plt, CRP and ESR were positively correlated. In addition, Plt positively correlated with Leu and ESR negatively correlated with DB and HSA. The non-enzymatic blood antioxidants, DB and HAS, correlated negatively with only ESR, RDW (DB only), and CRP (HSA only) (Table 3).

## Discussion

Cancer ranks as a leading cause of death and an important barrier to increasing life expectancy in every country of the world (57). Furthermore, it implies a burden not only for patients (due to disability and, therefore, the loss of working capacity) but also for society (caregivers, and family members). Hence the fundamental importance of valid preventive interventions, even before an early diagnosis. In this scenario, redox phenomena represent a common trait of almost all cancer hallmarks recognized so far. Conditions of oxidative stress are associated not only with the onset and progression of tumors but also with neoplastic treatment and it is often unavoidable side or unwanted effects. Therefore, having objective biochemical information on the current functioning of the redox system is a great challenge of modern oncology. Biochemical tests, such as the d-ROMs test and the BAP test, based on the use of photometers, the most common tools of any laboratory, have proved useful in providing a fairly reliable idea of the redox status of cancer patients. To our memory, this is the first study performed with the aim of evaluating the d-ROMs test and the BAP test in a population of cancer patients in Brazil.

Numerous studies have demonstrated the usefulness and effectiveness of the evaluation of the redox state through these two tests, both in the evaluation in humans and in animals (11,14,36,42,43,47,58–60) however, this is the first time that there was a correlation investigation of both parameters with a wide range of hematological and biochemical variables.

The evaluation of the redox state in the present study identified that the d-ROMs values were above normal levels (420.2 ± 112.1 U CARR), while the BAP test value was within the levels considered adequate (2332 ± 812 μmol/l). In addition, blood markers of inflammation, CRP, RDW, and Plt correlated positively with d-ROMs, OSI and negatively with BAP/dROMs (except the RDW).

ROS metabolism in cancer cells is an area of research that is still in progress, but it is already well established that an excess of ROS is involved in the process of carcinogenesis, both as risk factors and in the progression of the disease (10). Elevated serum levels of d-ROMs have already been identified in other studies involving individuals with various pathologies including cancer (34,42,43,46,49,61,62). Like in the meta-analysis performed by GÀO et al. (2019), who identified high levels of d-ROMs in cancer patients and a direct association of this parameter with the risk of developing lung, breast, and colorectal cancer (11). Another study that found high levels of d-ROMs in cancer patients was developed by MONTOVANI et al.; (2002) in this work, the authors used d-ROMs levels to monitor the impact of an antioxidant therapy on the management of oxidative stress (63).

Increased reactive oxygen species in cancer patients, as observed in the present study, can be caused by various tumor progression mechanisms, such as hypoxia, tumor cell survival, proliferation, chemo- and radio-resistance, invasion, angiogenesis and metastasis (64,65) and may act, and contribute to a series of pathophysiological events directly related to disease progression. Established events include involvement in cell cycle progression and proliferation, apoptosis, cell adhesion, motility, and tumor maintenance (4,10,18).

The analysis of comparison between the genders revealed the d-ROMs was significantly higher in women. This result agrees with other studies carried out in healthy individuals (47) and individuals with various pathologies (35,43,46,49,59,66). One of the possible explanations for this result is the hormonal changes in women associated with blood concentrations of estrogen and, therefore, on menopausal status (46).

The present study sought to investigate associations between oxidative stress markers and blood parameters. In this sense, important correlations were found, such as the positive correlation between the d-ROMs and BAP parameters, proving the close relationship between the pro-oxidant/ antioxidant system.

Another important result was the positive correlations found between d-ROMs, OSI, RDW, Plt, CRP and ERS, confirming an interrelationship between oxidative stress and inflammation parameters. This relationship is complex, but several studies prove the close association between this process (5,15,60,67) including in cancer (4,12,68). In the present study, a direct association was found between the oxidative parameters d-ROMs and OSI with the inflammatory markers RDW (d-ROMs only) (69), platelets (70,71), ESR and CRP (72).

Since it has been demonstrated that a strong, positive, and independent association exists between RDW and conventional inflammatory biomarkers (73) several studies have been developed and have confirmed RDW as an interesting variable for inflammatory predictor in various disorders including functional bowel conditions, autoimmune diseases, malignancy, COVID-19 and multiple hospital admissions in subjects with chronic conditions (69). In agreement with the present study, Karakilcik et al., (2014) and Semba et al., (2010) demonstrated a close association between RDW and biomarkers of oxidative stress. The reason for this relationship is the fact that oxidative stress exerts a profound effect on red blood cell homeostasis, such as a change in size, increasing its heterogeneity and therefore RDW (74,75). Therefore, oxidative stress increased red cell turnover, thus contributing to the association between anisocytosis and human pathology (69).

Another important finding of the study was the correlation between d-ROMs and platelets. This result indicates the complex relationship between oxidative stress, thrombotic processes and inflammation. Other studies have found similar evidence, such as the study developed by FRANCO et al (2015), who present an association between platelets, inflammation, and cancer (71). According to these authors, essential characteristics of cancer disease, previous described by Hanahan and Weiberg (2000) (76), such as cellular and microenvironment alterations necessary for malignant transformation, dysregulation of cell energetics, avoidance of immune destruction, genomic instability, and tumor-promoting inflammation resemble the inflamed state, placing the platelet within an interface that links thrombosis, inflammation, and cancer (71).

The ESR correlated positively with oxidative and inflammations markers, d-ROMs, OSI, Plt and CRP and negatively with antioxidants biomarkers, BAP/dROMs, DB and HAS. ESR is a non-specific index of inflammation which measures the rate at which red blood cells sediment in a period of one hour (72,77). The ESR and CRP are the most commonly used laboratory tests for detecting the acute phase response and thus diagnosis and monitoring of inflammatory condition. However, few studies have investigated the association of this parameter with oxidative stress. Some studies carried out with patients with rheumatoid arthritis (78) and depression (77) and showed that ESR to be affected by numerous physiologic and pathophysiologic conditions that involve not only changes of fibrinogen concentration in plasma but also alterations in the size, shape, and/or number of RBCs as well as presence of non–acute phase reaction proteins.

Other authors carried out investigations between the oxidative parameter d-ROMs with inflammatory variables CRP, such as FAIENZA et al. (2012) who found negative correlations between the BAP/d-ROMs index with CRP and cholesterol in obese children. FUKUI et al. (2011), found positive correlations between d-ROMs with CRP and creatinine in Japanese subjects. HIROSE et al., (2009), that found positive correlations between d-ROMs with CRP in diabetics individuals (79).

A large study developed by Aleksandrova et al., 2014 demonstrated through multivariable-adjusted logistic regression a positive association between d-ROMs/CRP with the risk of developing colorectal cancer (12). Excessive and uncontrollable production of reactive oxygen species results in persistent injury of cells in the tissue and consequently chronic inflammation (80). In turn, inflammatory cells produce soluble mediators, which act by further recruiting inflammatory cells to the site of injury and producing more reactive species. This sustained inflammatory/oxidative environment leads to an enhanced production of hydroperoxides in a vicious circle, which can damage healthy cells and over a long time may lead to carcinogenesis (67). In the case of patients already diagnosed with cancer, the high production of reactive oxygen species in preclinical tumors contributes to this cycle of oxy-inflammation.

The BAP/d-ROMs index correlated positively with the endogenous antioxidant’s albumin and bilirubin (borderline correlation) and negatively with platelet, CRP and ERS. The direct correlation between the index and the endogenous antioxidants, demonstrates this parameter is also useful to evaluate the relative antioxidant capacity. Furthermore, the inverse correlation with inflammatory variables corroborates the hypothesis of the close relationship between distress and inflammation.

The study had some limitations such as the sample size, being a retrospective study in which it was not possible to consider lifestyle factors, tumor severity or treatments. Despite this, important results with scientific basis were found, concluding that through the analysis of the d-ROMs and BAP tests, the redox state of cancer patients is in a situation of potential oxidative stress, in which despite the reactive species being produced in excess, the antioxidant system is working to achieve redox balance. Furthermore, hematological and inflammatory markers are parameters that directly affect the redox state and can influence to the development of oxidative distress.

The results found suggest that the d-ROMs and the BAP tests are simple and reliable for the diagnosis of oxidative stress and were associated with markers of inflammation. The measurement of these parameters provides information about oxidation and inflammation condition and can have important implications both in monitoring the health status of patients, the efficacy and safety of the treatments.

## Data Availability

All relevant data are within the manuscript and its Supporting Information files.

## Author Contributions

Conceptualization, Clarissa Aires de Oliveira, Eugenio Luigi Iorio, Foued SalmenEspindola; Methodology Eugenio Luigi Iorio and Clarissa Aires de Oliveira, Lara Ferreira Paraiso; Data curation Lara Ferreira Paraiso — original draft preparation, Clarissa Aires de Oliveira, Eugenio Luigi Iorio, Lara Ferreira Paraiso and Foued Salmen Espindola; writing—review and editing, Foued Salmen Espindola and Eugenio Luigi Iorio; visualization, Foued Salmen Espindola; supervision, Foued Salmen Espindola; project administration, Foued Salmen Espindola and Clarissa Aires de Oliveira; funding acquisition, Food Salmen Espindola and Clarissa Aires de Oliveira. All authors have read and agreed to the published version of the manuscript.

